# SARS-CoV-2 Seroprevalence among First Responders in the District of Columbia, May – July 2020

**DOI:** 10.1101/2020.11.25.20225490

**Authors:** Jacqueline Reuben, Adrienne Sherman, James A. Ellison, Jayleen K. L. Gunn, Anthony Tran, Matthew McCarroll, Pushker Raj, Patricia Lloyd, Preetha Iyengar, Fern Johnson-Clarke, John Davies-Cole, LaQuandra Nesbitt

**Author notes:** **Correspondence for DC Health:** Jacqueline Reuben, DC Health. **Correspondence for CDC:** James Ellison, Centers for Disease Control and Prevention.

## Abstract

First responders are at increased risk of occupational exposure to SARS-CoV-2 while providing frontline support to communities during the COVID-19 pandemic. In the District of Columbia (DC), first responders were among the first people exposed to and infected with SARS-CoV-2, with over 200 first responders diagnosed with COVID-19 by May 15, 2020. From June – July 2020, DC Health conducted a serologic survey to estimate SARS-CoV-2 seroprevalence and assess risk factors and occupational exposures among a convenience sample of first responders in DC. Of the 310 first responders tested, 3.5% (n = 11) had anti-SARS-CoV-2 antibodies. Seropositivity varied by occupation, with 4.8% (3/62) of firefighters; 3.6% (8/220) of police officers; and no paramedics (0/10) or administration and support staff (0/18) testing positive. Type and consistency of personal protective equipment (PPE) use also varied: all paramedics (n=10) reported wearing a N95 respirator all or most of the time, compared to 83.3% of firefighters, 38.8% of police officers, and 23.5% of administration and support staff (p<0.001). All paramedics reported wearing gloves all or most of the time, compared to 80.0% of firefighters, 27.8% of administration and support staff, and 24.3% of police (p<0.001). The relatively low seroprevalence among first responders highlights the benefits of continuous training on and reinforcement of the proper use of PPE while performing job duties to mitigate potential transmission within and between first responders and the community.

**Summary:** Understanding occupational exposure to and infection with SARS-CoV-2 among first responders is important for workforce planning and emergency preparedness and response. Seroprevalence among first responders (3.5%; 11/310) who participated in a survey conducted by the District of Columbia Department of Health (DC Health) from May 28 – July 15 was 48% lower than reported in the DC community (6.7%; 876/12990). The lower prevalence of SARS-CoV-2 among first responders highlights the importance of continuous training on and reinforcement of the proper use of personal protective equipment (PPE). Proper use of PPE is a critical mitigation strategy to reduce transmission among and between first responders and the community.

## Background

First responders, including personnel in law enforcement, fire and rescue, and emergency medical response, are on the frontlines of providing support and medical services to communities during the COVID-19 pandemic and are crucial to the public health and medical response. This close contact with community members, and with other first responders, places them at increased risk of occupational exposure to SARS-CoV-2 [1]. Proper use of personal protective equipment (PPE) is an important layer of defense that first responders have to protect themselves and to prevent the spread of COVID-19 infection [2, 3]. This may be especially true when higher level controls to occupational exposure are not feasible in the specific work setting or situation. Understanding how first responders utilize available PPE ensures sufficient resources, training and attention are given to support first responders in safely providing essential services, while mitigating community spread of COVID-19.

Early cases of COVID-19 infection in the District of Columbia (DC) were identified among first responders, with 104 fire and emergency medical services (FEMS) staff and 116 Metropolitan Police Department (MPD) staff diagnosed by nucleic acid testing between March 7 – May 15, 2020 [4]. It is not known, however, how many first responders have developed antibodies to SARS-CoV-2, and their use of PPE has not been described. The aims of this paper are to estimate the prevalence of seropositivity (i.e., presence of antibodies to SARS-CoV-2) and assess self-reported PPE use by occupation type among first responders in DC.

## Methods

### Study Design and Participant Enrollment

DC Health and the DC Department of Forensic Sciences Public Health Laboratory (DFS-PHL) partnered with Providence Health System, the District of Columbia Fire and Emergency Medical Services Department (DC FEMS), and the Metropolitan Police Department of the District of Columbia (DC MPD) to conduct a cross-sectional study to estimate the seroprevalence of SARS-CoV-2 among first responders, and assess risk factors and occupational exposures. Enrollment took place from May 18 – July 5, 2020. Participating agencies were responsible for recruiting their staff. DC FEMS distributed a mass email communication to the 1,934 operational members of their workforce informing them of this initiative and inviting them to participate. To recruit DC MPD staff, the Chief of Police emailed the 4,488 active MPD employees on four separate occasions to inform staff and encourage all to participate.

First responders were defined as any employee, staff, or volunteer working for or contracted by the District to perform fire safety and rescue, police, or any other function to protect the health, safety, and welfare of the public. This included personnel in law enforcement, fire and rescue, and emergency medical response. Only those serving as first responders in the District were eligible for inclusion. Otherwise eligible persons who reported having a positive COVID-19 diagnostic test within the past 14 days or reported onset of symptoms consistent with COVID-19 in the past seven days of questionnaire administration were excluded. All participants provided informed consent. This activity was determined to meet the requirements of public health surveillance as defined in 45 CFR 46.102(l).

### Questionnaire

Prior to serologic testing, participants were provided a link to complete an electronic questionnaire eliciting information on demographics, potential COVID-19 exposures, PPE use, medical history, and recent symptoms (Appendix A). Participants were asked to recall PPE use at work (while within 6 feet of a suspected or confirmed case of COVID-19), potential exposures, and symptom history dating back to March 1, 2020. Questionnaire data were collected and managed using the secure Research Electronic Data Capture (REDCap) application. Following completion of the questionnaire, eligible participants were asked to schedule an appointment for serologic testing at Providence Health System, a local urgent care facility located in Washington, DC.

### Serologic Testing

Serum specimens were collected by healthcare providers employed by Providence Health System. Samples were centrifuged within one hour of collection, refrigerated, and transported to the DC DFS-PHL for testing. All testing was completed within 48 hours of specimen receipt. Serologic testing was conducted using the DiaSorin LIAISON® XL assay (DiaSorin Inc., Stillwater, MN), a qualitative chemiluminescent assay for determination of immunoglobulin G (IgG) antibodies to the spike protein (anti-S1 and anti-S2) of SARS-CoV-2. The assay has been determined to be highly sensitive (97%) for specimens collected over 14 days post positive diagnostic test (DiaSorin IFU) [5]. The assay was used under an Emergency Use Authorization (EUA) for COVID-19 *in vitro* diagnostics issued by the United States Food and Drug Administration (FDA).

### Statistical analysis

The final analytic sample included only those who completed both the questionnaire and serology testing. Descriptive analyses were performed on the overall sample and reported by serological test result and by occupational group.

Proportions were reported for categorical variables, and differences by seropositivity were tested using 2-tailed *χ*^2^ tests for association with significance defined at the 0.05 level. Statistical analyses were performed using SAS software (version 9.4; SAS Institute).

## Results

Between May 28 – July 15, 2020, a total of 399 eligible first responders completed the online questionnaire, of whom 310 (77.7%) also completed serology testing. No statistically significant differences were observed between those who completed the questionnaire only, and those who completed both serology testing and the questionnaire. Among the 310 first responders tested, 3.5% (n = 11) had anti-SARS-COV-2 antibodies (Table 1). Seropositivity varied by self-reported occupation, with 4.8% (3/62) of firefighters, 3.6% (8/220) of police officers, no (0/10) paramedics, and no (0/18) administration and support staff testing positive. The mean age of participants was 42.5 years (range: 22–68 years) and 76.5% (n=237) were male. The majority of participants reported identifying as White (53.6%; n=166) or Black/African American (29.4%; n=91). Nine percent (9%; n=28) identified as Hispanic/Latino. Self-reported Hispanic ethnicity was significantly associated with seropositivity (p=0.04). Over half of participants (52.9%; n=164) were residents of Maryland. No persons with SARS-COV-2 antibodies indicated a pre-existing medical condition (Table 1). As shown in Table 2, 64.8% (n=201) of participants had no prior SARS-CoV-2 test via nasal, throat or saliva sample. Testing behavior varied by serology test result with 45.5% (n=5) of seropositive participants reporting no previous test, compared to 65.6% (n=196) of seronegative participants. The most frequently reported symptoms (since March 1, 2020) among those who tested positive were: headache (72.7%; n=8), cough (63.6%; n=7), fever (54.6%; n=6), chills (54.6%; n=6), sore throat (54.6%; n=6), and muscle ache (54.6%; n=6). Seropositive first responders reported more frequent contact with patients with COVID-19, with 85.7% (n=6) of seropositive first responders reporting exposure to a known positive, compared to 33.8% (n=49) of seronegative responders (p<0.01). More seropositive participants reported having a positive household member than seronegative participants (57.1% vs 4.0%, respectively; p<0.01).

**Table 1:** Characteristics of first responders who obtained testing for COVID-19 antibodies between May 28 – July 15, 2020 in Washington DC.

| Demographics | Antibody Negative<br>N (%)<sup>a</sup> or Mean (SD) | Antibody Positive<br>N (%)<sup>a</sup> or Mean (SD) | Total<br>N (%)<sup>a</sup> or Mean (SD) |
| --- | --- | --- | --- |
| Total | 299 | 11 | 310 |
| Age, years (SD) | 42.60 (8.96) | 41.09 (12.21) | 42.55 (9.07) |
| Sex at Birth |  |  |  |
| Female | 69 (23.08) | 4 (36.36) | 73 (23.55) |
| Male | 230 (76.92) | 7 (63.64) | 237 (76.45) |
| Race |  |  |  |
| White | 162 (54.18) | 4 (36.36) | 166 (53.55) |
| Black/African American | 87 (29.10) | 4 (36.36) | 91 (29.35) |
| Asian | 11 (3.68) | 0 (0.0) | 11 (3.55) |
| American Indian/Alaska Native | 2 (0.67) | 0 (0.0) | 2 (0.65) |
| Native Hawaiian/Pacific Islander | 2 (0.67) | 1 (9.09) | 3 (0.97) |
| Other/Multi-racial | 20 (6.69) | 2 (18.18) | 22 (7.10) |
| Declined to Answer | 15 (5.02) | 0 (0.0) | 15 (4.84) |
| Ethnicity<sup>*</sup> |  |  |  |
| Hispanic/Latino | 25 (8.80) | 3 (27.27) | 28 (9.49) |
| Non-Hispanic/Non-Latino | 259 (91.20) | 8 (72.73) | 267 (90.51) |
| State of Residence |  |  |  |
| District of Columbia | 84 (28.09) | 3 (27.27) | 87 (28.06) |
| Maryland | 158 (52.84) | 6 (54.55) | 164 (52.90) |
| Virginia | 52 (17.39) | 1 (9.09) | 53 (17.10) |
| Other | 5 (1.67) | 1 (9.09) | 6 (1.94) |
| Occupation |  |  |  |
| Police Office | 212 (70.90) | 8 (72.73) | 220 (70.97) |
| Firefighter | 59 (19.73) | 3 (27.27) | 62 (20.00) |
| Paramedic | 10 (3.34) | 0 (0.0) | 10 (3.23) |
| Other<sup>b</sup> | 18 (6.02) | 0 (0.0) | 18 (5.81) |

<sup>\*</sup> p<0.05 <sup>a</sup> Numbers may not sum to total due to missing values. Percentages are column percentages. <sup>b</sup> Other: administration and support staff

| Self-Reported Medical History | Negative<br>N (%)<sup>a</sup> | Positive<br>N (%)<sup>a</sup> | Total<br>N (%)<sup>a</sup> |
| --- | --- | --- | --- |
| Total | 299 | 11 | 310 |
| Diabetes |  |  |  |
| Yes | 15 (5.15) | 0 (0.0) | 15 (4.97) |
| No | 276 (94.85) | 11 (100.0) | 287 (95.03) |
| Hypertension |  |  |  |
| Yes | 43 (14.78) | 0 (0.0) | 43 (14.24) |
| No | 248 (85.22) | 11 (100.0) | 259 (85.76) |
| Chronic Heart Disease |  |  |  |
| Yes | 0 (0.0) | 0 (0.0) | 0 (0.0) |
| No | 287 (100.0) | 11 (100.0) | 298 (100.0) |
| Chronic Kidney Disease |  |  |  |
| Yes | 2 (0.69) | 0 (0.0) | 2 (0.67) |
| No | 287 (99.31) | 11 (100.0) | 298 (99.33) |
| Chronic Liver Disease |  |  |  |
| Yes | 0 (0.0) | 0 (0.0) | 0 (0.0) |
| No | 289 (100.0) | 11 (100.0) | 300 (100.0) |
| Asthma |  |  |  |
| Yes | 23 (8.01) | 0 (0.0) | 23 (7.72) |
| No | 264 (91.99) | 11 (100.0) | 275 (92.28) |
| COPD<sup>c</sup> |  |  |  |
| Yes | 2 (0.69) | 0 (0.0) | 2 (0.67) |
| No | 286 (99.31) | 11 (100.0) | 297 (99.33) |
| Immunosuppressed Condition |  |  |  |
| Yes | 4 (1.38) | 0 (0.0) | 4 (1.33) |
| No | 285 (98.62) | 11 (100.0) | 296 (98.67) |

COPD = Chronic Obstructive Pulmonary Disease
<sup>a</sup> Numbers may not sum to total due to missing values. Percentages are column percentages.
<sup>b</sup> Other: administration and support staff

**Table 2:** COVID-19 testing behavior, symptoms experienced, and exposure related information of first responders between May 28 – July 15, 2020 in Washington DC.

|  | Negative<br>N (%)<sup>a</sup> | Positive<br>N (%)<sup>a</sup> | Total<br>N (%)<sup>a</sup> |
| --- | --- | --- | --- |
| Total | 299 | 11 | 310 |
| Number of Times Previously Tested |  |  |  |
| 0 | 196 (65.55) | 5 (45.45) | 201 (64.84) |
| ≥ 1 | 103 (34.45) | 6 (54.55) | 109 (35.16) |
| Fever<sup>*</sup> |  |  |  |
| Yes | 40 (14.8) | 6 (60.00) | 46 (15.75) |
| No | 242 (85.82) | 4 (40.40) | 246 (84.25) |
| Chills<sup>*</sup> |  |  |  |
| Yes | 32 (11.43) | 6 (54.55) | 38 (13.06) |
| No | 248 (88.57) | 5 (45.45) | 253 (86.94) |
| Cough<sup>*</sup> |  |  |  |
| Yes | 57 (20.14) | 7 (63.64) | 64 (21.77) |
| No | 226 (79.86) | 4 (36.36) | 230 (78.23) |
| Sore Throat<sup>*</sup> |  |  |  |
| Yes | 57 (20.28) | 6 (54.55) | 63 (21.58) |
| No | 224 (79.72) | 5 (45.45) | 229 (78.42) |
| Shortness of Breath<sup>*</sup> |  |  |  |
| Yes | 23 (8.10) | 5 (45.45) | 28 (9.49) |
| No | 261 (91.90) | 6 (54.55) | 267 (90.51) |
| Vomiting |  |  |  |
| Yes | 6 (2.17) | 0 (0.00) | 6 (2.10) |
| No | 270 (97.83) | 10 (100.00) | 280 (97.90) |
| Diarrhea |  |  |  |
| Yes | 47 (16.85) | 3 (27.27) | 50 (17.24) |
| No | 232 (81.15) | 8 (72.73) | 240 (82.76) |
| Muscle Aches<sup>*</sup> |  |  |  |
| Yes | 61 (21.79) | 6 (54.55) | 67 (23.02) |
| No | 219 (78.21) | 5 (45.45) | 224 (76.98) |
| Loss of Smell or Taste<sup>*</sup> |  |  |  |
| Yes | 6 (1.17) | 2 (20.00) | 8 (2.79) |
| No | 271 (97.83) | 8 (80.00) | 279 (97.21) |
| Headache<sup>*</sup> |  |  |  |
| Yes | 75 (26.60) | 8 (72.73) | 83 (28.33) |
| No | 207 (73.40) | 3 (27.27) | 210 (71.67) |
| Positive Coworker |  |  |  |
| Yes | 97 (52.15) | 6 (75.00) | 103 (53.09) |
| No | 89 (47.85) | 2 (25.00) | 91 (46.91) |
| Positive Household Member <sup>*</sup> |  |  |  |
| Yes | 10 (3.95) | 4 (36.36) | 14 (5.30) |
| No | 243 (96.05) | 7 (63.67) | 250 (94.70) |
| Other Known Positive Individuals <sup>*</sup> |  |  |  |
| Yes | 49 (33.79) | 6 (85.71) | 55 (36.18) |
| No | 96 (66.21) | 1 (14.29) | 97 (63.82) |

\*p<0.01
<sup>a</sup> Numbers may not sum to total due to missing values. Percentages are column percentages.

The most frequently reported use of any PPE (Table 3) was an N95 respirator, with 49.2% reporting wearing a respirator all or most of the time, followed by 38.6% (n=112) reporting using gloves all or most of the time. Gown and goggle/face shield use while on duty was lower, with 75.9% (n=221) of participants reporting never or rarely wearing a gown, and 70.9% (n=197) reporting never or rarely using goggles or a face shield. While there were no statistically significant differences in PPE use by serology test result, seropositive participants reported more consistent use of a respirator other than an N95 or surgical mask, with 18.2% (n=2) of seropositive individuals reporting use all or most of the time, compared to 3.9% (n=10) of seronegative individuals (p=0.07).

**Table 3:** Personal Protective Equipment (PPE) use among first responders by serology outcome between May 28 – July 15, 2020 in Washington DC.

|  | Negative<br>N (%)<sup>a</sup> | Positive<br>N (%)<sup>a</sup> | Total<br>N (%)<sup>a</sup> |
| --- | --- | --- | --- |
| Total | 299 | 11 | 310 |
| N95 Respirator |  |  |  |
| All/most of the time | 139 (49.12) | 5 (50.00) | 144 (49.15) |
| Sometimes | 65 (22.97) | 2 (20.00) | 67 (22.87) |
| Rarely/Never | 79 (27.92) | 3 (30.00) | 82 (27.99) |
| Surgical Mask |  |  |  |
| All/most of the time | 99 (36.00) | 4 (36.36) | 103 (36.01) |
| Sometimes | 70 (25.45) | 2 (18.18) | 72 (25.17) |
| Rarely/Never | 106 (38.55) | 5 (45.45) | 111 (38.81) |
| Goggles/Face Shield |  |  |  |
| All/most of the time | 61 (22.85) | 2 (18.18) | 63 (22.66) |
| Sometimes | 18 (6.74) | 0 (0.00) | 18 (6.47) |
| Rarely/Never | 188 (70.41) | 9 (81.82) | 197 (70.86) |
| Gloves |  |  |  |
| All/most of the time | 108 (38.71) | 4 (36.36) | 112 (38.62) |
| Sometimes | 80 (28.67) | 4 (36.36) | 84 (28.97) |
| Rarely/Never | 91 (32.62) | 3 (27.27) | 94 (32.41) |
| Gown |  |  |  |
| All/most of the time | 44 (16.48) | 2 (18.18) | 46 (16.55) |
| Sometimes | 21 (7.87) | 0 (0.0) | 21 (7.55) |
| Rarely/Never | 202 (75.66) | 9 (81.82) | 211 (75.90) |

<sup>a</sup> Numbers may not sum to total due to missing values. Percentages are column percentages.

PPE use varied significantly by occupation (Table 4). All paramedics (100%; n=10), 83.3% (n=50) of firefighters and 38.8% (n=80) of police officers indicated that they wore an N95 respirator all or most of the time (p<0.01). Firefighters and paramedics reported more consistent gown use, with 61.7% (n=37) of firefighters and 50% (n=5) of paramedics reporting wearing a gown all or most of the time, compared to 2.1% (n=4) of police officers (p<0.01). Paramedics and firefighters reported more consistent glove use, with 100% (n=10) of paramedics and 80% (n=48) of firefighters reporting use all or most of the time, compared to 24.3% (n=49) of police officers. Goggle/face shield use varied across occupation, with 100% (n=10) of paramedics, 75% (n=45) of firefighters and only 4.2% (n=8) of police officers indicating they wore goggles/face shields all or most of the time.

**Table 4:** Personal Protective Equipment (PPE) by occupation type between May 28 – July 15, 2020 in Washington DC.

|  | Police Officer<br>N (%)<sup>a</sup> | Firefighter<br>N (%)<sup>a</sup> | Paramedic<br>N (%)<sup>a</sup> | Other<br>N (%)<sup>a, b</sup> | Total<br>N (%)<sup>a</sup> |
| --- | --- | --- | --- | --- | --- |
| Total | 220 | 62 | 10 | 18 | 310 |
| N95 Respirator<sup>*</sup> |  |  |  |  |  |
| All/most of the time | 80 (38.83) | 50 (83.33) | 10 (100.0) | 4 (23.53) | 144 (49.15) |
| Sometimes | 60 (29.13) | 3 (5.00) | 0 (0.0) | 4 (23.53) | 67 (22.87) |
| Rarely/Never | 66 (32.04) | 7 (11.67) | 0 (0.0) | 9 (52.94) | 82 (27.99) |
| Surgical Mask<sup>*</sup> |  |  |  |  |  |
| All/most of the time | 70 (34.83) | 21 (36.21) | 2 (22.22) | 10 (55.56) | 103 (36.01) |
| Sometimes | 64 (31.84) | 5 (8.62) | 0 (0.00) | 3 (16.67) | 72 (25.17) |
| Rarely/Never | 67 (33.33) | 32 (55.17) | 7 (77.78) | 5 (27.78) | 111 (38.81) |
| Goggles/Face Shield<sup>*</sup> |  |  |  |  |  |
| All/most of the time | 8 (4.19) | 45 (75.00) | 10 (100.0) | 0 (0.0) | 63 (22.66) |
| Sometimes | 14 (7.33) | 4 (6.67) | 0 (0.0) | 0 (0.0) | 18 (6.47) |
| Rarely/Never | 169 (88.48) | 11 (18.33) | 0 (0.0) | 17 (100.0) | 197 (70.86) |
| Gloves<sup>*</sup> |  |  |  |  |  |
| All/most of the time | 49 (24.26) | 48 (80.00) | 10 (100.0) | 5 (27.78) | 112 (38.62) |
| Sometimes | 73 (36.14) | 5 (8.33) | 0 (0.0) | 6 (33.33) | 84 (28.97) |
| Rarely/Never | 80 (39.60) | 7 (11.67) | 0 (0.0) | 7 (38.89) | 94 (32.41) |
| Gown<sup>*</sup> |  |  |  |  |  |
| All/most of the time | 4 (2.08) | 37 (61.67) | 5 (50.00) | 0 (0.0) | 46 (16.55) |
| Sometimes | 10 (5.21) | 7 (11.67) | 4 (40.00) | 0 (0.0) | 21 (7.55) |
| Rarely/Never | 178 (92.17) | 16 (26.67) | 1 (10.00) | 16 (100.0) | 211 (75.90) |

<sup>\*</sup>p<0.001
<sup>a</sup> Numbers may not sum to total due to missing values. Percentages are column percentages.
<sup>b</sup> Other: administration and support staff

## Discussion

In a convenience sample of 310 first responders in the District, 3.5% (n=11) had detectable levels of IgG SARS-CoV-2 antibodies at the time of serologic testing. Initial estimates among U.S. healthcare personnel have suggested that frontline healthcare workers (HCWs) account for 10–20% of all diagnoses [6]. These reports, however, did not differentiate by specific occupation type or potential source of exposure. We found that seroprevalence among first responders in DC (3.5%) was 48% lower than the seroprevalence in the general population (6.7%) tested by the DC PHL during this time period. The seropositivity observed in this sample of first responders was comparable to the seroprevalence of SARS-CoV-2 IgG antibodies observed in a sample of 203 firefighters/paramedics in South Florida (3.9%) [7]. No prior studies, however, have reported risk among law enforcement, although they may interface closely with communities where ongoing transmission from asymptomatic individuals has been documented [8].

At the time of survey, FEMS PPE policies required members to wear a N95 respirator, gloves, and face shield for every patient encounter within six feet, and an N95 respirator, gloves, face shield, gown, bouffant head covering, and shoe coverings when encountering any patient with confirmed or suspected COVID-19. MPD PPE policies required all members to wear a cloth or surgical mask while on duty, and a minimum of an N95 respirator and gloves when encountering any patient with confirmed or suspected COVID-19 or responding to any medical emergency call. Both FEMS and MPD reported having no shortages of PPE supplies during this time. Per DC FEMS and DC MPD protocols, all occupations should consistently wear PPE that is appropriate for the context of their exposures. CDC guidelines for first responders recommend at minimum use of an N95 respirator or face mask when interacting with a suspected or confirmed patient with COVID-19 [1]. Consistent use of N95 respirators and surgical masks among this sample of first responders may account for the relatively low seropositivity compared to the general population. Use of certain PPE varied by occupation, as different types of PPE may be necessary to perform required duties and guidance on use varies by occupation. This survey found that among police officers, less than half reported wearing an N95 respirator all or most of the time, and less than 25% reported wearing gloves all or most of the time while at work and within six feet of a person with suspected or confirmed COVID-19. A study of EMS encounters with patients with COVID-19 found that only 0.4% of providers tested positive during the 14 days following an encounter when 67% of EMS used full PPE including mask, eye protection, gown, and gloves [9]. These results suggest that continual training on the importance of consistent and proper PPE use may be needed among first responders to ensure protocols continue to be followed correctly over time.

This study was limited by its non-random convenience-based sampling strategy. Given the low response rate and small sample size, particularly among paramedics, these estimates may not be generalizable to the entire first responder workforce in DC. Retrospective self-report of exposures and PPE use is subject to bias, and PPE availability was not assessed through the questionnaire. The study population was limited to active members of the DC MPD and FEMS workforces and excluded any individuals with suspected active COVID-19 infection as indicated by a positive PCR test in the past 14 days or onset of COVID-like symptoms in the past seven days.

Despite these limitations, to our knowledge this is one of the first SARS-CoV-2 seroprevalence surveys conducted specifically among a cohort of first responders including law enforcement, thus providing valuable information to the agencies responding to the pandemic in the District of Columbia and adding to the limited literature surrounding PPE use and SARS-CoV-2 seroprevalence in this population. In recognition of the important role that face coverings play in reducing the spread of COVID-19, DC Mayor Bowser issued Mayor’s Order 2020-080 on July 22, 2020, mandating face masks or face coverings to be worn outside the home by all individuals in the District. This mandate requires not only DC residents, but also visitors and employees (inclusive of first responders) working in the District to wear a face covering at all times when not at home.

## Conclusion

Estimating the seroprevalence of COVID-19 among first responders helps inform adequate emergency preparedness and workforce safety. First responders are at high risk for acquiring infections during novel disease outbreaks. The relatively high proportion of participants reporting using N95 respirators or surgical face masks may help explain the lower seropositivity despite the high risk of occupational exposure. While recommended PPE use in CDC guidelines varies by occupation and setting, the significant differences in PPE use across occupation types in this study suggests the need to assess consistent use of PPE in the context of an ongoing pandemic [1]. Additional training on the importance of wearing appropriate PPE while performing job duties may be needed to ensure workforce safety and mitigate the spread of COVID-19.

## Data Availability

All data is provided in table format within the manuscript file.

## Acknowledgements

The authors would like to thank the invaluable contributions and assistance of members of the CDC EOC and DC Health COVID-19 response teams, and laboratorians at the DC DFS-PHL who have worked tirelessly to ensure prompt and accurate test results, as well as Dr. Senai Medhani, Dr. Shadi Soufi and their clinical team at Providence Health System for their support and assistance in the collection of serum specimens.

## Disclaimer

Use of trade names and commercial sources are for identification only and do not imply endorsement by the US Department of Health and Human Services nor the Government of the District of Columbia. The findings and conclusions in this report are those of the authors and do not necessarily represent the views of their institutions.

## Supplemental Information

https://coronavirus.dc.gov/maskorder

https://www.acludc.org/en/letter-mayor-bowser-requesting-mandatory-use-face-masks-all-dc-police-officers

https://www.cdc.gov/coronavirus/2019-ncov/community/organizations/firefighter-EMS.html

https://www.cdc.gov/coronavirus/2019-ncov/community/guidance-law-enforcement.html

https://www.cdc.gov/niosh/topics/hierarchy/default.html

https://www.diasorin.com/sites/default/files/allegati/liaisonr_sars-cov-2_s1s2_igg_brochure.pdf.pdf

